# The impacts of artificial intelligence on the workload of diagnostic radiology services: A rapid review and stakeholder contextualisation

**DOI:** 10.1101/2025.07.24.25332132

**Authors:** Claire Sutton, Julie Prowse, Mai Elshehaly, Rebecca Randell

## Abstract

**Background:** Advancements in imaging technology, alongside increasing longevity and co-morbidities, have led to heightened demand for diagnostic radiology services. However, there is a shortfall in radiology and radiography staff to acquire, read and report on such imaging examinations. Artificial intelligence (AI) has been identified, notably by AI developers, as a potential solution to impact positively the workload of radiology services for diagnostics to address this staffing shortfall.

**Methods:** A rapid review complemented with data from interviews with UK radiology service stakeholders was undertaken. ArXiv, Cochrane Library, Embase, Medline and Scopus databases were searched for publications in English published between 2007 and 2022. Following screening 110 full texts were included. Interviews with 15 radiology service managers, clinicians and academics were carried out between May and September 2022.

**Results:** Most literature was published in 2021 and 2022 with a distinct focus on AI for diagnostics of lung and chest disease (n = 25) notably COVID-19 and respiratory system cancers, closely followed by AI for breast screening (n = 23). AI contribution to streamline the workload of radiology services was categorised as ‘autonomous’, ‘augmentative’ and ‘assistive’ contributions. However, percentage estimates, of workload reduction, varied considerably with the most significant reduction identified in national screening programmes. AI was also recognised as aiding radiology services through providing second opinion, assisting in prioritisation of images for reading and improved quantification in diagnostics. Stakeholders saw AI as having the potential to *remove some of the laborious work* and contribute service resilience.

**Conclusions:** This review has shown there is limited data on real-world experiences from radiology services for the implementation of AI in clinical production. Autonomous, augmentative and assistive AI can, as noted in the article, decrease workload and aid reading and reporting, however the governance surrounding these advancements lags.

## Introduction

Globally the work of radiology services is extensive [1]. Increasingly volume and task complexity for radiologists has grown [2, 3]. Whilst technical advances in medical imaging benefit patient outcomes, most increase the workload for diagnostic radiology [3]. Ageing populations, limitations on training posts in radiology and the impacts of COVID-19 have exacerbated a global shortage of radiologists [4].

Hosny et al. [5^501^] advised that artificial intelligence (AI) ‘… within the imaging workflow would increase efficiency, reduce errors and achieve objectives with minimal manual input’. Gampala et al.’s [6] review recommended AI proffers radiologists more time for both patient consultation and complex image analysis. The potential of AI to impact radiology services positively was advised by the European Society of Radiology (ESR) [7]. An ESR [8^2^] survey of members in December 2018 identified in general a ‘favourable attitude’ to AI in radiology. However there continues to be scepticism regarding AI in clinical radiology. Wong et al. [9^142^] reported fear ‘computers replace radiologists’ but advised AI has the potential to change clinical radiology for advancement.

Karantanas and Efremidis [10] advocated for radiologists to ‘control’ the application and prevalence of AI in clinical practice. Eltawil et al.’s [11^1^] scoping review examining factors shaping facilitators and barriers advised of radiologists’ ‘lack of acceptance and trust’ of AI as well as its potentiality for improved accuracy and efficiency. Eche et al. [12] noted limitations in generalisability of AI attenuates adoption at pace in clinical practice. Similarly, Gisanti [13] has noted the need for robust, consistent, governance with respect to AI adoption particularly with an international lens.

Machine learning (ML) was one of the first terms used to describe computers learning as they were exposed to datasets [7]. The evolution of ML has led to complex models that aim to simulate human thinking that in turn has led to deep learning algorithms of many layers of connected artificial neural networks capable of complex tasks such as image interpretation [7]. Computer-aided detection/diagnosis (CAD) according to Fujita [14] heralded a third wave of AI development.

Categorisation of CAD is by clinical application for example: detection, diagnosis and triage with implementation often for prioritisation purposes [14]. Whilst the emphasis of computer-aided diagnostics is on accuracy, sensitivity and specificity, for detection there has been a recent call for studies that look at the impact on, ‘clinically meaningful endpoints such as survival, symptoms, and need for treatment’ [15^e486^].

This rapid review of the impacts of AI on the workload of radiology services was initially undertaken as part of a workforce review of radiology services in the north of England. The workforce review also comprised a series of interviews with radiology services’ stakeholders who were asked specifically about AI. This rapid review and contextualising interviews with UK radiology service stakeholders aimed to synthesise a range of research to respond to the following question pertaining to the impacts of AI on radiology services workload: *What and how, does and could artificial intelligence impact radiology services’ workload now or in the future?*

## Materials and Methods

A rapid review, noted as an accelerated means of evidence synthesis [16, 17], was selected as an appropriate review approach, within a five-month timeframe (April to August 2022), to harness and interpret available and appropriate international evidence pertinent to the research question.

Guidance from the Cochrane Rapid Reviews Methods Group [18] was used to structure the rapid review protocol. Once the protocol was agreed between the review team it was lodged with the ‘International prospective register of systematic reviews’ (PROSPERO) and is identifiable by the reference: CRD42022341257.

The following five databases were searched: arXiv, Cochrane Library, Embase, Medline and Scopus with variations of the following search string used in Embase, Medline and Scopus: “radiolog*” AND (“artificial intelligence” OR “machine learning” OR “deep learning” OR “image analysis”) AND

(“workload” OR “skill mix” OR “productivity”). Computer science and engineering databases were not incorporated owing to the focus of the review being the impact of AI on health service delivery rather than technological advancement of AI. Searches were limited by English language literature published between January 2007 and May 2022. These databases and date range were chosen to provide a comprehensive level of literature without unnecessary duplication. A health studies subject librarian provided expert guidance regarding literature searching. Search results from across the five databases were collated into a single EndNote library. Following deduplication, the title and abstract of each record were screened. Subsequently the full texts of all retrieved records were screened. The inclusion and exclusion criteria utilised to develop a full text screening algorithm are provided in Table 1.

**Table 1:** Inclusion and exclusion criteria.

| Inclusion criteria | Exclusion criteria |
| --- | --- |
| Published in English | Published in a language other than English |
| Published between 01 January 2005 and 31 December 2022 | Published before 01 January 2005 and after 31 December 2022 |
| Studies reporting empirical data pertaining to:<br>• AI in clinical radiology for diagnostics in human healthcare<br>• How AI impacts clinical radiology workload<br>• How AI could change the skill mix/productivity of clinical radiology services | Literature reviews, conference proceedings or posters without empirical data or full accounts or empirical data, Master's or PhD thesis, book chapters or books that may have reported:<br>• AI in clinical radiology for diagnostics in human healthcare<br>• How AI impacts clinical radiology workload<br>• How AI could change the skill mix/productivity of clinical radiology services |
| Policy paper about AI impact/s on clinical radiology workload | Any other political paper about AI impact/s on clinical radiology workload or related area |

The flowchart for full text screening, which was developed from the initial title/abstract screening algorithm, is presented in Appendix 1. The Mixed Methods Appraisal Tool (MMAT) [19] was used to evaluate the quality of included literature (see Appendix 3). Data extraction culminating in thematic analysis and narrative synthesis of key findings was completed manually by one author. The production of the rapid review by a single researcher was a divergence from the protocol lodged with PROSPERO however was necessary owing to limited time to complete the rapid review initially. The findings of this review are contextualised with quotes from interviews with UK radiology service stakeholders including senior clinicians, managers and academics involved in the education of radiographers. The interview data reported in this paper is focused on AI in clinical diagnostic radiology services and consequences for the workforce, however the whole interview schedule, of which the AI oriented question was a part, was more far-ranging.

Our choice to contextualise a rapid review with stakeholder views aligns to Christensen et al.s [20] suggestion, if review findings are to be impactful, their implementation might be enhanced through the clinicians’ ‘voices’. Greenhalgh and Papoutsi [21^2^] have previously advocated for the importance of research that engages and represents the complexity of healthcare, ‘we need research designs and methods that foreground dynamic interactions and emergency’.

If our approach aligns to Braun and Clarke’s [22] small q research, then we propose, despite a non-representative sample of stakeholders, lack of data saturation and triangulation, the ‘voices’ of senior clinicians, managers and academics do provide legitimate insights into how AI is viewed on the ‘frontline’ of clinical diagnostic radiology services. We used purposive sampling to identify fifteen interviewees who, as Johnson, Adkins and Chauvin [23] advise, had specific experiences and expertise we wanted to learn about in relation to clinical diagnostic radiology services in the UK. The following AI specific question was asked to service managers and clinicians: *What are your thoughts about introducing artificial intelligence into radiology services?* And to academics: *What do you think is the potential for digital technology, for example artificial intelligence, to support cancer diagnosis within radiology and what are the consequences in terms of workforce planning?*

Interviews were carried out between May and September 2022 and recorded using Microsoft Teams. Recordings were transcribed verbatim by a commercial transcription service and manually analysed using Braun and Clarke’s [24] approach to thematic analysis. Ethical approval for the empirical work was granted by the Research Ethics Committee of the University of Bradford (reference: EC27051).

## Results

### 1) Overview of included literature

Figure 1 provides an overview of the literature selection process of the review. A summary in tabular format of the included literature (n=110) is presented in Appendix 2. Results of the quality appraisal undertaken of each item of included literature are presented in Appendix 3.

**Figure 1:**
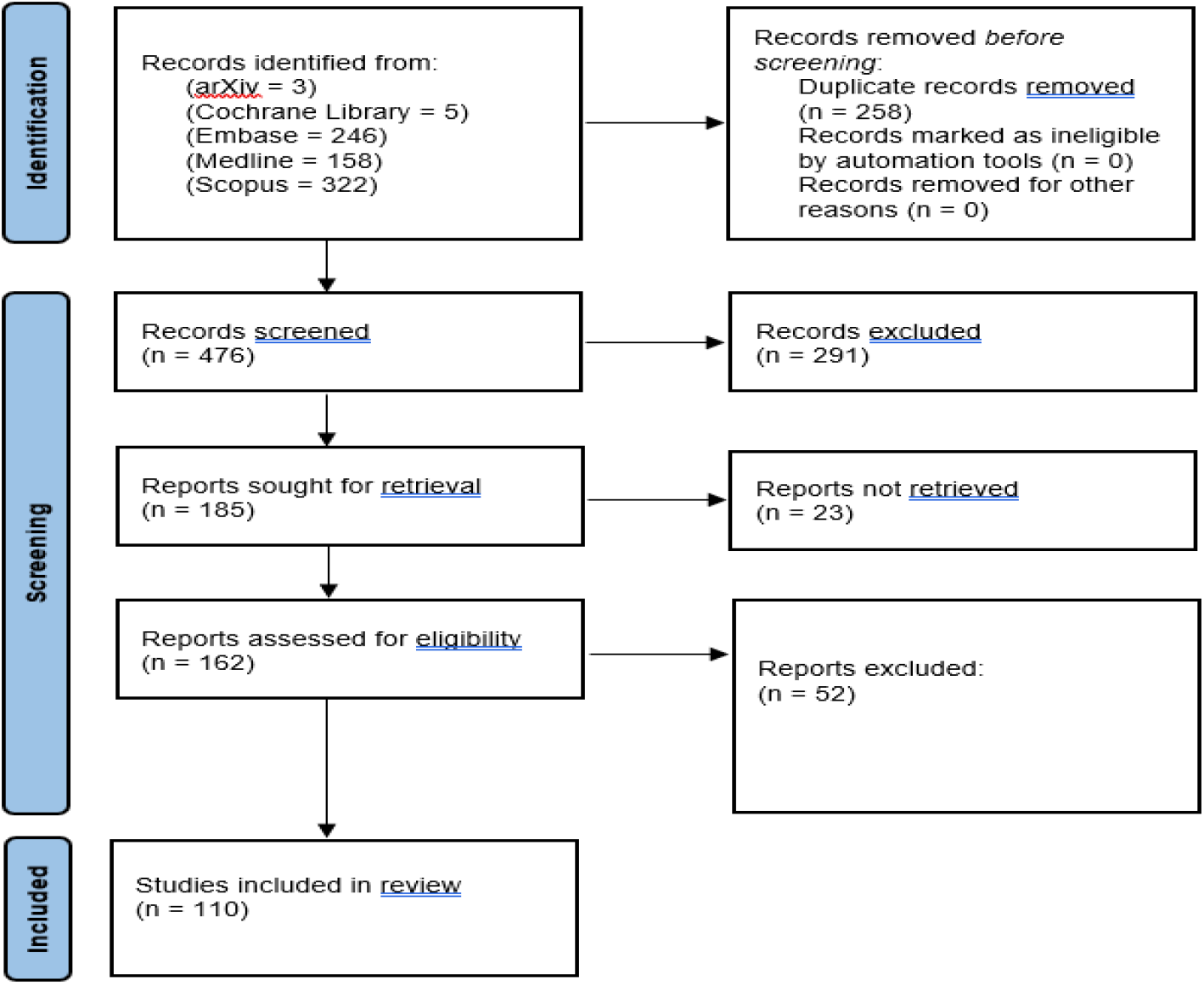
Simple flow chart to show numbers of articles at each stage of the selection process

The greatest diversity in quality across the body of literature of the review occurs with the implementation of statistical analysis (question 4.5). The variation in sample size for both AI training and evaluation is also notable, with one research project using as few as 50 X-rays [25] and another, scans from 114 421 participants [26].

Most literature included within the review was published in 2021 (n = 35) with a noticeable increase in publications from 2020 onwards. The bar chart below (Figure 2) illustrates included literature of the review by year.

**Figure 2:**
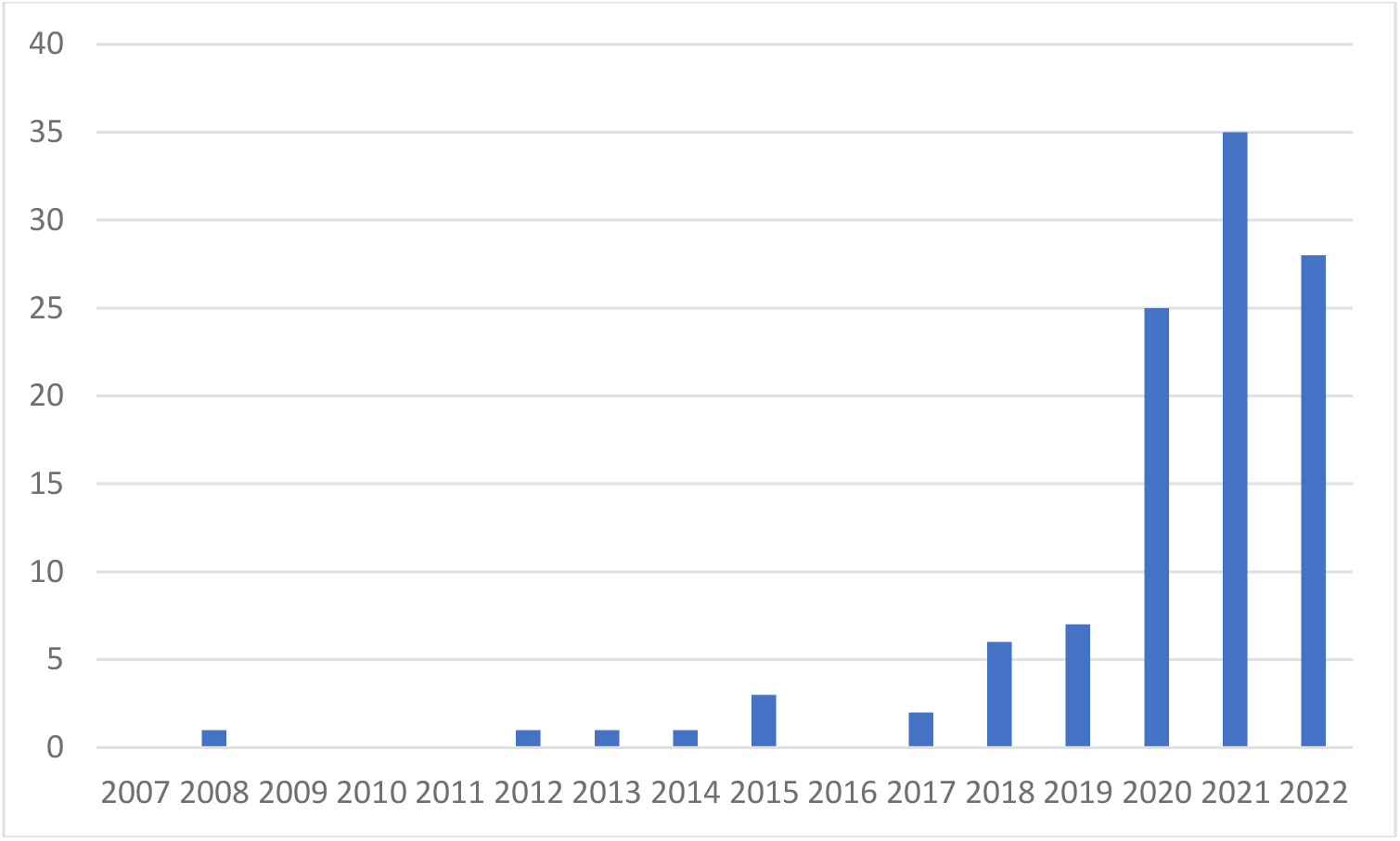
Included literature by year from January 2007 to May 2022

There is a global distribution of included literature. Most literature is from researchers in China (n = 33), followed by 13 items of literature from researchers in the United States (US), eight items of literature from international research collaborations and six items of literature from researchers based in India.

It is apparent researchers often relied on or incorporated publicly available datasets to develop and evaluate AI, for example those identified by Agrawal and Choudhary [27] including Imagenet; Japanese Society of Radiological Technology database; from Shenzhen No. 3 People’s Hospital in China; Department of Health and Human Services Montgomery County, Maryland; and the National Institutes of Health Chest X-ray Dataset. Similarly, Alduraibi [28] utilised the Breast Ultrasound Images Dataset. Hidayah et al. [29] used the University of California Irvine Machine Learning Repository. The Lung Image Database Consortium (LIDC-IDRI) was used in the work of Ismaeil and Salem [30]; Kasinathan and Jayakumar [31]; Monkam et al. [32]; Seidel et al. [33]; Tandungan et al. [34] and Zamacona et al. [35].

References to AI for diagnostics of lung and chest disease most frequently occur, with n = 25. Twenty-three items of literature examined AI for COVID-19 diagnosis and pneumonia secondary to COVID-19. Similarly, 23 items of literature presented AI for breast screening; pathologies including cancer. AI application in imaging of the central nervous system is illustrated in eight items and seven items focus on acute ischaemic stroke, haemorrhage infarction and intracranial aneurysms.

Notably AI in cancer diagnostics is apparent in 50 studies with most concerned with breast cancer (n = 24), lung nodules, diseases including cancer (n = 17) and cancers of the central nervous system (n = 3). All, except one item of literature, referenced the impacts of AI for the workload/workflow of medics by the terms ‘doctors’, ‘radiologists’, ‘physicians’, ‘medical practitioners.’ Pedrosa et al. [36] referenced the impact of AI for ‘technicians’ and ‘radiologists.’ On occasion medical specialities are referenced for example as ‘nuclear medicine physicians’ [37] and ‘neuroradiologists’ [38].

Computed Tomography (CT) is the most common imaging examination utilised for AI development and evaluation (n = 39). Radiographs are also used extensively. Most radiographs were chest X-rays (n = 21). Magnetic Resonance Imaging (MRI) and Magnetic Resonance Angiography (MRA) are referenced as the basis for AI development and evaluation in 19 studies. Mammography was the imaging examination of choice in 13 studies.

The most commonly described AI in the review’s literature is a type of deep learning: convolutional neural networks (CNNs) (n = 45). However, there is variation in how authors described CNNs in their research [37, 39–42]. ‘Deep learning’ is also frequently referenced (n = 17). Computer-aided diagnosis/detection (CAD) including utilisation of image segmentation is represented as a common application (n = 27) in the literature often from CNNs. CAD is used for disease detection and in disease classification for diagnosis often coupled with image segmentation (n = 7). Automating image segmentation with, for example, weakly supervised CAD would aid physician workload reduction considerably [43, 44]. The contribution of AI to automate segmentation is further discussed in the next section. Machine learning is referenced in a small number of studies (n = 5).

Radiomics is also referenced in a small number of studies (n = 4) benefiting quantifiable data interpretation notably in the diagnosis of lung nodules [45], pancreatic cancer detection and/or classification [46] and glioma grading [47].

### 2) Artificial intelligence (AI) for reducing volume of workload and improving service efficiency

Percentage estimates for reductions in clinical radiologists’ workload owing to AI implementation are variable, from 15% [48] to more than 86% [26]. Greatest estimates, notably of up to 86.7% [26, 49] in clinical radiologists’ workload, are seen in the context of national screening programmes [26, 45, 50]. This evidence is echoed in the words of a regional director for health and social care workforce transformation who stated:

> *The technology that’s out there now should be reading 90% of what needs reading … the really complicated (images) should only be read (manually) …*

This review notes categories of possible AI contribution and clinician oversight termed ‘independent’, ‘semi-dependent’ and ‘dependent’ to categorise how AI may positively impact radiology workload and efficiency. AI-human interaction, as determined in Allen’s [51] discussion, in relation to the regulation of AI in pathology, has been informative. Allen [51] categorised the contribution of AI as ‘in-the-loop’, ‘on-the-loop’ and ‘out-of-the-loop’ with ‘the loop’ being the level of the clinician’s responsibility for the AI contribution. Whilst the focus of this review is not primarily the level of clinician responsibility and thus supervision of AI, from a legislative perspective, level of clinician responsibility is helpful to group the ways in which AI may positively reduce radiology workload and enhance service efficiency. Table 2, below, provides explanations of this review’s categories of ‘autonomous’, ‘augmentative’ and ‘assistive’ AI and an overview of radiology workload of these categories of AI contribution to reduce radiology workload and aid service efficiency.

**Table 2:** Categories of AI contribution to reduce radiology workload and enhance service efficiency.

| Category of AI contribution | Radiology workload impacts |
| --- | --- |
| Autonomous AI: AI reads and reports an image and may offer a patient management plan independently of clinician input | National screening programmes to exclude 'without disease', 'normal' or 'non-positive' reports; potentiality for use in settings without an expert clinician to read, report and proffer patient management plan |
| Augmentative AI: AI reads and reports an image that a clinician may then utilise in further patient management planning | Automation of segmentation to reduce clinician cognitive strain; offer supplementary or auxiliary diagnosis; provide second opinion with benefits for less experienced clinicians |
| Assistive AI: AI reads an image, the data of which is then utilised by a clinician in their reporting and further patient management planning | Improved sensitivity and specificity particularly for quantifying lesions to aid manual reading; reduce reading time (in comparison with sole manual reading time) |

#### i) Autonomous AI

AI has the potential to automate fully initial steps in radiology diagnostics notably through differentiating imaging examinations by those ‘with’ from those ‘without’ disease; termed computer-aided detection or diagnosis. AI, particularly utilising convolutional neural networks, for the automated detection of COVID-19 on chest X-rays for rapid disease identification is reported widely in the literature [42, 52, 53, 54, 55]. Similarly automated detection of ‘normal’ from ‘abnormal’ mammograms is of increasing interest [56]. The words of a general manager for radiology services of a large acute healthcare provider are illustrative of this category of AI:

> *(We’re) starting a project quite soon in breast imaging … we’re looking at whether AI … on the PACS system, will be able to interpret, and then just one radiologist or one reporting radiographer (will report) … it’ll supplement the current workforce or remove some of the laborious work, or the work that’s maybe deemed not necessary. So, it might provide some extra resilience as well*.

Efficiency of triage, the process by which image/patient management may be sequenced, can be aided by AI application. For illustration, the work of Galvan-Tejada et al. [73^14^] advised their AI ‘may have the practical use of triaging mammograms in developing countries where there is a deficiency of expert readers.’ Rodriguez-Ruiz et al. [83^4830^] proposed AI ‘to automatically pre-select exams for radiologist evaluation …’. This thought chimes with the words of a clinical professor in radiography:

> *I think the key (thing) from our perspective is the work that’s going on at the moment around second read, around prioritisation of work. So, (AI) can triage, and certainly this is one of those questions we’re having currently within our pathways … (for example) a chest x-ray with suspicious findings, it will do an immediate read, and it will put it to the top of the worklist*.

Verburg et al. [84] and Yala et al. [85] similarly proposed AI for automated triaging breast screening notably for benefit where double reading by clinicians is standardised practice (in Europe) to aid clinical efficiency; AI plus clinician compared with two clinicians. Rajaraman et al.’s [65^28^] AI for tuberculosis classification proffered ‘advanced assistance in radiologist interpretive workflows to include triage of abnormal findings …’. Zapaishchykova et al. [86^424^] proposed AI, based on classification, for ‘an automated yet interpretable pelvic trauma decision support system’.

#### ii) Augmentative AI

One of the most significant means by which AI positively impacts radiology workload is automation in segmentation, this is referenced in more than 20 items of literature of the review. Segmentation is a process for image analysis, often coupled with classification, where segmentation owing to image sub-division leads to refined tissue contouring, and classification aids image level disease prediction based on image data [27]. Convolutional neural networking is utilised commonly for AI driven automation in segmentation [39, 40, 57, 58, 59, 60, 61, 62, 63, 64, 65, 66, 67, 68].

Particularly, interest is shown in the use of AI to automate lung segmentation in the detection and/or diagnosis of COVID-19 [39, 68, 69], lung disease [65, 71] and lung nodules [58]. Similar interest is also evident in AI for automation in segmentation of known or suspected malignancy for example in the analysis of images of breast lesions [40, 60, 71], hepatocellular carcinoma [57] and nasopharyngeal carcinoma [60]. Sichtermann et al.’s [66] AI to automate segmentation of intracranial aneurysms offered the potential of automation in segmentation to reduce radiologists’ cognitive strain. Van der Oever et al.’s [67] AI for detection of coronary artery calcium (CAC) exclusion or segmentation benefited a reduction in 86% of all scans for CAC requiring radiologist review.

AI can also contribute to radiology workload efficiency by acting as a tool for supplementary diagnosis. Chen et al. [72^231^] evaluated deep transfer learning, for rapid detection and classification for COVID-19, claiming improved sensitivity and reduced reading time versus manual reading concluding transfer learning as a model for ‘auxiliary diagnosis’. Similarly, multiple researchers termed AI as a tool for ‘second opinion’. Ali et al. [70] advised AI proffering second opinion increases objectivity in diagnosis. Galvan-Tejada et al. [73] noted AI for second opinion supports classification of breast tumours. Hussain et al. [76] advised their AI gave radiologists a second opinion during interpretation to aid COVID-19 diagnosis. Noshad et al. [42] noted AI may provide second opinion not only for decreased workload but improved accuracy in COVID-19 detection. Comparatively Pedrosa et al.’s [36^15^] research presented a more sceptical view of deep learning performances for COVID-19 screening however nonetheless did conclude AI could provide a ‘robust’ second opinion.

AI has the potential to support less experienced radiologists as an adjunctive diagnostic tool. Kakeda et al. [38] identified AI benefits improved accuracy in the reading of MR angiography for intracranial haemorrhage by less experienced radiologists; however less experienced radiologists plus a computer-aided diagnosis system were no more accurate than experienced neuroradiologists. In addition, those less experienced radiologists assisted by a computer-aided diagnosis system (CADS) were quicker in their reading than experienced neuroradiologists without CADS. Yao et al. [82] in the context of rib fracture detection also concluded AI aids clinician reporting performance including speed of diagnosis for both junior and experienced radiologists.

Rao et al. [77^92^] recommended utilising AI in the context of intracranial haemorrhage diagnosis ‘as an adjunct to current peer review tools as a second reader …’. Wang et al.’s [78] work also in relation to intracranial haemorrhage (detection and classification), proposed AI for second reading or triaging; workflow facilitation. AI for supplementary diagnosis or ‘second opinion’ may streamline interpretation and thus reduce the number of clinicians to read and report. McKinney et al. [77^93^] noted AI for breast cancer screening has the potential to reduce ‘the need for double reading in 88% of UK screening cases …’ with no loss of accuracy. A general manager for radiology services of a large healthcare organisation advised of planned augmentative AI trialling noting:

> *… (in) …. asymptomatic breast screening, where they’re looking for … early identification of cancers … historically, they’ve always had two people, but it could be that (this is reduced to one) … I think that will really support … we have a significant workforce challenge for radiologists and consultant radiographers in the breast area*.

Zamacona et al. [35] utilised AI for diagnostic categorisation and determination of subsequent radiologist reading requirement. Grauhan et al. [87^355^] promoted AI for worklist prioritisation and ‘safety in situations of increased workload.’ A divisional director for clinical support services in a large healthcare organisation advised in relation to augmentative AI:

> *… I’m sure it’ll [AI] be enormous in the future. … whether it’s in assisting reporting … (or) supporting decision making around (reporting)*.

#### iii) Assistive AI

AI has been found to reduce the time taken to read individual scans. Benedikt et al. [80] concluded their computer-aided detection system with digital breast tomosynthesis can reduce reading time by 29.2%. Joshi et al. [54] concluded their AI is capable of detecting COVID-19, from non-COVID-19 on chest X-rays in 0.137 seconds. Li et al. [61] claimed their AI for automatic segmentation of imaging in total anomalous pulmonary venous connection produced results in 400 milliseconds compared with two to three hours by manual segmentation. Improved rapidity of disease detection or diagnosis may expedite patient management, notably treatment, particularly in urgent cases but may also improve communication between clinicians. For example, Meng et al. [81] reported AI for timely communication between radiologists and referring physicians where the AI was able to identify, based on radiological findings, those cases requiring prompt follow-up. A consultant radiologist noted their positivity about the possibilities of assistive AI although felt it would particularly benefit timely data consensus:

> *I am fully positive about it. (Imagine) … if you started a report and you had all that information, like, “Lesion X is this, the lung nodules measure this …” and a comparison to a previous (series of images) … and then you just had to join it all up, look for the extra findings … that would massively speed up our reporting*.

AI may aid improved sensitivity and specificity in delineating areas of suspicion, notably suspected malignancies. Antonios et al. [71] described BreDAn to automate detection, differentiation and staging of breast lesions. Chang et al. [86^1^] concluded their AI was capable of ‘reliable quantitative lung diagnosis …’. AI can also benefit improved sensitivity in the detection of small, asymptomatic, vascular lesions. For example, Cao et al. [43^1^] reported AI to detect acute ischaemic stroke and haemorrhagic infarction finding AI ‘had significantly greater patient-level sensitivity than did the human readers.’ A consultant radiologist expressed their expectations for assistive AI:

> *I hope (AI) makes (us) better image interpreters, whether radiographers or radiologists … (however) maybe the only thing I’d love it to do for me is to measure lesions in cancer*.

Increasingly AI becomes shared knowledge for international implementation, and available for commercial distribution. For example, both Johansson et al. [89] and Lauritzen et al. [26] evaluated the application of Transpara® as a computer-aided detection/diagnosis system in mammography. Aiello et al. [69] evaluated a range of AI-based segmentation tools for the diagnosis of COVID-19.

Similarly, Ardakani et al. [90] appraised 10 convolutional neural networks for the management of COVID-19 diagnosis. Applications of ResNet, a residual neural network which can be used for image classification, are referenced in the work of Grauhan et al. [85]; Hseih et al. [37]; Jacob et al. [89] and Zhang and Zhang [68]. U-Net, a convolutional neural network for image segmentation is referenced in the work Gaal et al. [58]; Khaled et al. [61]; Liu et al. [92]; Mahmood et al. [63]; Rajaraman et al. [65]; Zhang and Zhang [68]. It is worth noting all research literature reviewed involved AI development and/or evaluation with the impacts for workload/workflow noted as secondary outcomes.

## Discussion

There is a growing body of international evidence on AI’s potential to reduce the volume and improve the efficiency of manual labour required to be undertaken by radiology services. A decreased volume of images for manual reading is likely to reduce radiologists’ reading fatigue and facilitate focus of clinicians’ attention on high-risk, complex reading tasks, benefiting not only clinician accuracy but also potentially improved patient interactions. In addition, AI can aid prioritisation of the reading queue aiding workflow efficiency. However, there is no consensus that AI used concurrently with radiologist interpretations, such as ‘autonomous’ or ‘augmentative’, excluding segmentation tools, accounting for most of the legally marketed AI tools in the US and Europe, improves radiology service efficiency. AI may not necessarily improve radiology services efficiency given the increased resources required to evaluate and/or adjudicate AI inference. There is limited primary research examining the real-world adoption of AI and impacts on radiology service workload and efficiency.

There is heterogeneity across the included studies of this review. Whilst most studies of this review focus on AI development and/or evaluation, sources of datasets, types of AI and anticipated contributions of AI are diverse. Pigott and Shepperd’s [69] classification of sources of heterogeneity is useful here since it advocates for consideration in how interventions have been implemented, context and conduct of studies notably in complex interventions – those of clinical application in real-world settings. Pigott and Shepperd’s [69] characteristics of heterogeneity are also evident in Yu et al.’s [70] contemporary study that examined predictors of heterogeneous treatment effects of AI assistance in diagnostic performance of radiologists.

AI design, with the use of deep learning or convolutional neural networks for computer-aided detection and/or diagnosis, and performance evaluation, are the foci of the literature consulted although the scale of such AI design and evaluation research is variable. AI to reduce the volume of work, particularly across large national programmes, for example in breast screening [84, 85] and thoracics [8] develops globally, yet implementation of AI and subsequent patient management around the world is variable [56]. At present there is a limited literature that specifically evaluates radiology services’ efficiencies, clinician uptake and patient outcomes of AI implementation.

Reddy et al.’s [93] review of AI evaluation frameworks called for evaluation to run in parallel with AI development and workflow integration. This review echoes this call for more research to establish real world impacts of AI on radiology workload and workflow. Vasey et al. [94] have proposed a systematised approach to clinical evaluation of AI for ‘bringing AI systems from mathematical performance to clinical utility’. The majority of the studies of this review sought to evaluate the accuracy of an AI product with possible impacts on clinician workload and workflow as assumptive conclusions.

Whilst stakeholder comments presented within this review indicated positive anticipation of AI’s capability to reduce workload, aid reading prioritisation, improve reading accuracy as well as proffer remote supervision of more junior staff by their senior colleagues, consistent adoption and implementation of AI for radiology diagnostics is more nuanced. Stakeholders’ positivity chimes with the findings of Scott et al. [95] and Chew and Achananuparp [96], however does not reflect the concerns also noted in the literature relating to patient privacy breaches and liability for AI errors raising concerns about trustworthiness of AI-led results. Knop et al.’s [97] work has highlighted the need to better understand human-AI interactions notably from professionals’ perspectives. A recent review by Cartolovni et al. [98^1^] concluded AI for clinical decision-making shifted a ‘bilateral patient-physician relationship … into a trilateral one’ with at present limited consideration for the authenticity of such dimensions. Similarly Young et al.’s [99] work examining patient and public views on AI found general positivity towards AI however also presented patients’ caution and predilection for human-interaction. AI can contribute a solution to improve workflow efficiency of radiology services, although the extent to which it can make such a contribution will be determined by the ongoing debate regarding the ultimate legal responsibility for particularly autonomous AI. For example, a recent letter from the US Food & Drug Administration [100] advised on the use of an AI tool for radiology, stating that images not flagged by the system with suspected findings, which are potentially negative cases, still need to be interpreted, raising the question of whether the use of AI to screen out negative cases in cancer screening programmes is a realistic prospect.

### Strengths and limitations of the review

A single author retrieved, screened and extracted data from literature for the review. This was not planned for within the review protocol (CRD42022341257), however it is not believed to have led to bias within the review and of significant detriment to the quality of the review’s findings. Ultimately a single author approach to the review aided the speed at which the review was produced initially to meet the timeframe stipulated. However, this said, all interviews, quotation from which have been utilised to contextualise specific findings of the literature review, were carried out by the second author. The combination of findings from both a rapid review with interview data is a less common research approach compared with a standalone review, or standalone interviews, however such a combination is believed to strengthen the findings from the rapid review alone carried out by a single author. Whilst the stakeholder contextualisation may be identified as subjective and biased towards the benefits of AI adoption notably among radiology services in the UK, nevertheless, the real-world views presented do illustrate the need for further empirical work to evaluate more rigorously and extensively clinicians’ experiences of AI with respect to workload and flow in this fledging area of research.

## Conclusion

There has been and is a global ambition to develop AI, with the benefit to reduce the workload for clinicians of diagnostic radiology services, particularly since 2020. Most studies, of this review, centre the use of AI in imaging for the diagnosis of lung and chest disease, and coincide with diagnostic demands associated with the COVID-19 pandemic. In the future, the use of AI to increase clinicians’ time for complex reading tasks may have greatest impact in national screening programmes, notably through autonomous AI. Autonomous AI reads and reports, with a patient management plan, independently of clinician contribution. This may be compared with assistive AI that reads and reports data for the clinician to utilise in patient management, and augmentative AI reads and provide data for the clinician to use in their reporting and patient management. Despite the potential of AI to reduce clinicians’ workload and promote workflow efficiency in diagnostic radiology services, there is very little empirical research that evaluates or adjudicates AI inference, similarly examines the real-world adoption of AI and its impacts. There are significant concerns from clinicians for patient privacy and confidentiality, and legislative responsibility for the use of AI in clinical diagnostic radiology services globally.

## Data Availability

All data produced in the present work are contained in the manuscript.

## Acknowledgements

This paper is based on commissioned research undertaken by the Workforce Observatory, that was funded by the West Yorkshire Integrated Care Board on behalf of the West Yorkshire Health and Care Partnership. We thank the interview participants for contributing their time and perspectives.

## Appendices

1. Flowchart for full text screening
2. Summary of included literature following full text screening
3. Quality appraisal of included literature using the Mixed Methods Appraisal Tool (2018)

