## Supplemental file for "The impacts of artificial intelligence on the workload of diagnostic radiology services: A rapid review and stakeholder contextualisation"

**Appendices**

A) Flowchart for full text screening
B) Summary of included literature following full text screening
C) Quality appraisal of included literature using Mixed Methods Appraisal Tool (2018)

**Appendix A Flowchart for full text screening**

Where possible the full texts of all ‘included’ records following title/abstract screening have been attached to the title/abstract record in an EndNote library. Where full texts are not available (from UoB library or the inter-library loan request system) for any record, the abbreviation: NA may be added to the ‘Research Notes’ for the record in the EndNote library; for example: 2, NA; 3, NA or 4, NA. For full texts that are not in English, the abbreviation: NE may be added into the ‘Research Notes’ of the EndNote library record, for example: 2, NE. Screening of available full texts may be achieved by using the following algorithm and updating the ‘Research Notes’ of the record in the EndNote library; for example: 2, 1.

English language study with empirical data about AI in clinical radiology for diagnostics in human healthcare and has a full text, published between 2005 and 2022/present, retrievable via UoB library/inter-library loans system that is not a literature review, conference proceeding or poster without empirical data/full account of empirical data, Master’s or PhD thesis, book chapter or book?

No

Yes/Maybe

Explains how AI impacts clinical radiology workload?

Yes/Maybe

No

Is a policy paper about AI impact/s on clinical radiology workload?

Explains how AI could change the skill mix/productivity of clinical radiology services?

Yes/Maybe

/?

No

No

Yes/Maybe

M

**Appendix B Summary of included literature following full text screening**

Total: 185 included papers following title/abstract screening; 110 included papers following full text screening and initial data extraction

| **Lead author (Surname) and Type of item** | **Date of publication and country of researchers/Research base** | **Full text screening code** | **How AI impacts radiology services’ workload** | **Notes (disease and radiology professionals)** |
| --- | --- | --- | --- | --- |
| Abbas, Full conference proceeding – development and evaluation of AI | 2020, Pakistan (50 X-rays) | 3, 2 | Transfer learning, faster R-CNN, for computer-aided diagnosis; automated fracture detection; X-rays | Lower limb fractures; Doctors/radiologists |
| Agrawal, Article – development and evaluation of AI: Efficient UNet | 2022, India with data from US (138 CXRs; 108 948 frontal CXRs) and from China (public dataset 662 images (normal lungs and those with tuberculosis) | 2, 2 | Possibility of automated diagnosis – detection and definition; segmentation; CXRs | Lung disease; Radiologists |
| Aiello, Article – evaluation of AI | 2022, Italy (validation dataset 73 CT images) | 2, 2 | Comparison of AI – segmentation tools; speed up diagnosis; CT images | COVID-19 (C-19) detection and monitoring; ‘supporting diagnosis and quantification … lung lesions.’ Radiologists |
| Alajmi, Article – development and evaluation of AI | 2022, Saudi Arabia | 3, 2 | Machine learning with image analysis for automated C-19 detection; CXRs | C-19 detection; Radiologists |
| Alduraibi, Article – development and evaluation of AI | 2022, Saudi Arabia (validation on 780 BU images – publicly available dataset) | 2, 3 | Convolutional neural networks and shallow classifiers; breast ultrasonography for diagnosis and differentiation; Enhance classification accuracy | Breast cancer (malignant vs. benign lesions); Radiologists/doctors |
| Ali, Full conference proceeding – development and evaluation of AI | 2020, US (AI tested on 566 CXRs in publicly available dataset) | 2, 3 | Computer-aided diagnosis/detection (CAD); lung segmentation; second opinion in reading, increase objectivity; CXR | Lung disease (C-19, lung cancer, tuberculosis, pneumonia) |
| Allaouzi, Full conference proceeding – development and evaluation of AI | 2021, Researchers in Morocco, international data sets (p. 60, example 5314 CXR images in one databased and dataset) | 2, 2 | Automated diagnosis; CXR | C-19 detection; Radiologists |
| Amara, Article – development and evaluation of AI | 2022, Algeria (50 scans from MedMod data set – Italian) | 3, 3 | Novel neural network architecture for automated segmentation; virtual reality platform for reading and visualisation support for better interpretation; CT | C-19 detection; Radiologists |
| Antonios, Article – development and evaluation of AI | 2013, Greece (BreDAn (clinical decision support system (CDSS)) clinical evaluated 534 MRI datasets; 3 case illustrations using BreDAn) | 2, 3 | Supplementation radiodiagnostic process, potential for automation with CDSS to CAD; captures exact pathological area through segmentation process; MRI | Breast pathology; Radiologists |
| Ardakani, Article – development and evaluation of AI | 2020, Iran, Singapore, Malaysia, Taiwan (10 CNNs on 108 patients with C-19; 89 patients with other pneumonias) | 3, 3 | 10 Convolutional neural networks (CNNs); RestNet-101 and Xception best performers as adjuvant tools detection C-19; CT | C-19 diagnosis; Radiologists |
| Benedikt, Article – evaluation of CAD on reading time | 2018, Data from France and the US (525 cases) | 3, 3 | Computer-aided detection in digital breast tomosynthesis; CAD with DBT leads to 29.2% faster reading time | Breast screening; Radiologists |
| Calisto, Article – an exploration of how clinicians interact with AI | 2021, Portugal (45 physicians across 9 clinical settings) | 3, 3 | Deep neural network (in human-centric AI) for automation and classification for workflow efficiency and quality; ‘high level of acceptance of AI .. reduction of cognitive workload … improvement in diagnosis’; Mammography | Breast imaging; Radiologists |
| Cao, Article – development and evaluation of weakly supervised learning | 2022, China (1027 patients) | 2, 3 | Weakly supervised learning /Diffusion-weighted imaging (MR imaging); location of small stroke lesions; more sensitivity than human; reduce labelling workload | Acute ischaemic stroke and haemorrhagic infarction lesions; Radiologists |
| Chang, Article – development and evaluation of multi-objective deep learning model | 2022, Taiwan (458 CT scans) | 3, 3 | Multi-objective deep learning; position, margin and texture lesions - quantitative lung diagnosis/report | Lung cancer; Radiologists |
| Chen, Article – AI development and evaluation (deep transfer learning) | 2021, China (422 patients) | 2, 3 | Deep transfer learning; recognition and classification C-19; less time than radiologists, auxiliary diagnosis; CT images | C-19; Radiologists |
| Chen, Article – AI development | 2021, China (3 data sets: ProMRI (gut MRI), ACDC (cardiology MRI) and REFUGE (retinal MRI) – international datasets) | 3, 3 | Weakly supervised segmentation; reduction in annotation costs; improved efficiency of delineation; MR images | Illustrative organ: prostate; Physicians |
| Clymer, Article – AI application | 2020, US | 2, 2 | Convolutional neural networks and transfer learning (CNNs); enhanced prioritisation; reduction in arthrograms – un-necessary invasive intervention; ‘second opinion’ (p. 5); MR images | Glenoid labrum/shoulder labral tear; Physicians; Radiologists |
| Cunha, Article – AI development and evaluation | 2020, US (484 patients) | 3, 3 | Segmentation and convolutional neural network (CNN); image quality evaluation and reduction in examination time – 48% examinations may have been reduced by applying CNN; MR images | Hepatocellular carcinoma screening/diagnosis; Radiologists |
| Dai, Article – development and evaluation of AI | 2021, China (265 patients, 17050 CT images for training and validation of p-EffNet) | 2, 3 | Deep learning/supervised convolutional neural network; enhanced detection; CT images | Arterial stenosis in lower limbs; Radiologists |
| Dembrower, Article – evaluation of AI for triage (commercial product) | 2020, Sweden (7364 women in study sample) | 2, 2 | AI cancer detector algorithm/deep neural network; AI as ‘single reader … no radiologist involvement, and to select women for enhanced supplemental reading … by radiologist’ (p. e472); Mammography | Breast cancer triage; Radiologists |
| Duron, Article – AI development and evaluation | 2021, France (600 patients; 60 170 X-rays: 70% training, 10% validation, 20% internal testing) | 3, 3 | Deep convolutional neural network; Improved fracture detection, time efficiency in localisation of fractures; X-rays | Skeletal fractures; Emergency physicians and radiologists |
| Dyer, Article – AI (for triage) evaluation | 2022, UK, US and India (390 CT head scans from UK and US for validation; 2059 CT scans for AI training) | 3, 3 | AI algorithm (EfficientNet neural network) with ground-truthing (to check the result of machine learning for accuracy); for AI to siphon abnormal scans to radiologist for expert reading, ‘AI as a decision support system’ (p. 740); CT scans | Intracranial haemorrhage, acute infarct; Ground-truth labelling by Consultant Radiologists (UK); AI for use in ED |
| Dyer, Article – evaluation of AI | 2021, UK (UK data) (3887 CXRs from 3790 patients) | 2, 3 | Deep-learning/convolutional neural network; Automated detection of normal CXRs (High Confidence Normal) – potential to remove 15% examinations from radiology reporting workflow; ‘DL … confidence scores and abnormal heatmaps (for non HCN)’ (p. 473e.14) | Diagnosis chest radiographs/CXRs; Radiologists |
| Feng, Full conference proceeding – AI development and evaluation | 2021, China | 3, 3 | Computer-aided diagnosis (CAD) for ‘diagnostic efficiency’, ‘for grading severity of knee OA’ (p. 3796); X-rays | Knee osteoarthritis; Radiologists |
| Gaal, Full conference proceeding – AI development and evaluation | 2020, Hungary (247 CXRs from Japanese Society of Radiological Technology; 138 CXRs from Montgomery; 662 CXRs from Shenzhen) | 3, 3 | Deep learning for organ segmentation; Fully convolutional networks; U-Net; CXRs | Chest X-rays in diagnostics for lung nodules, cardiothoracic ratio in cardiomegaly |
| Galvan-Tejada, Article – AI development and evaluation | 2017, Mexico (publicly available: Breast Cancer Digital Repository BCDR-D01: 64 women and BCDR-D02: 164) | 3, 3 | Computer-assisted diagnosis for classification of benign and malignant lesions, ‘detect abnormalities earlier’ (p. 2), ‘system .. potential … second opinion for … radiologist, or … practical use triaging mammograms’ (p. 14); Mammography/X-ray | Breast cancer; Radiologists |
| Ghosal, Article – AI development and evaluation | 2021, India (iSeg2017 dataset (US), 23 neonatal subjects and IBSR dataset (US), 18 MRIs from those 7 to 71 years) | 2, 2 | 3D deep learning, Multi headed U-Net for tissue/organ segmentation; AI differentiate: gray matter, white matter, CSF; MR images | Brain MR images; Radiologists |
| Grauhan, Article – AI development and evaluation | 2022, Germany (2700 shoulder radiographs for AI training) | 2, 3 | Convolutional neural network, ResNet-50; prioritisation of worklists; Radiographs/X-rays | Detect common causes of shoulder pain: fractures, joint dislocation, OA, periarticular calcifications, osteosynthesis and endoprosthesis; Physicians/clinicians |
| Hallinan, Article – AI development and evaluation | 2021, Singapore (100 lumbar spine ‘studies’/images) | 3, 3 | Deep learning, convolutional neural network, for classification tasks, ‘semi-automated reporting under the supervision of a radiologist to provide more consistent and objective reporting’ (p. 137); MR images | Detection/classification central canal, lateral recess and neural foraminal stenosis at lumbar spine MRI; Radiologists |
| Han, Article – AI development and evaluation | 2022, China (public dataset: 1018 CT scans from 1010 patients | 3, 3 | Computer aided diagnosis (CAD) using 3D CNN; ‘to help doctors pay more attention to those suspected early lung cancer nodules’ (p. 11); CT scans | Pulmonary nodules detection; Doctors |
| Hidayah, Full conference proceeding – AI development and evaluation | 2015, Indonesia (dataset from UCI Machine Learning Repository (US)) | 3, 3 | Machine learning in clinical decision-tree (J48) with classification; No description of initial dataset AI based | Detection and classification of vertebral column pathologies; Radiologists |
| Hsieh, Article – AI development and evaluation | 2021, China (37 427 sets of images of 19 041 patients at China Medical University Hospital) | 2, 3 | Deep learning, convolutional neural networks: ResNet and DenseNet; ‘help physicians confidently and safely rule out bone metastases’ (p. 11); whole-body bone scan | Bone metastases; Nuclear medicine physicians |
| Huang, Full conference proceeding – AI development and evaluation | 2019, China (3111 brain images (T1-C images) from Shengjing Hospital) | 2, 2 | Gamma correction and deep learning, convolutional neural network to train classifier; Automation of brain tumour screening; ‘radiologists … deal with … “tumors”’ (p. 52); MR images | Brain tumours; Radiologists |
| Hussain, Article – AI development and evaluation | 2022, Malaysia (images/X-rays for model development from publicly available database) | 3, 3 | Convolutional neural network; Automated detection C-19 with second opinion for clinicians; CXRs | C-19 detection; Radiologists |
| Ismaeil, Full conference proceeding – AI development and evaluation | 2020, Egypt (dataset from Lung Image Database Consortium dataset: 1018 thoracic CT scans with pulmonary nodules) | 3, 3 | Computer-aided detection, assistance in reading process; CT scans | Pulmonary nodules; Radiologists  EXCLUDE? – limited focus on how AI impacts workflow; focus is on sliding windows in AI tool, detection of pulmonary nodules in 2D slices c.f. to usual 3D slices |
| Jacob, Full conference proceeding – AI development and evaluation | 2021, India (888 CT scans from Lung Nodule Analysis challenge (US)) | 2, 2 | Convolutional neural network, pre-trained ResNet-50; automated detection pulmonary nodules (malignancy) | Pulmonary nodules (less than 3cm compared to pulmonary mass more than 3cm); Radiologists |
| Jang, Article – AI evaluation for C-19 detection (‘commercialized (deep learning) DL algorithm: Lunit INSIGHT for CR2’ (p. 3)) | 2020, Korea (279 patients) | 2, 3 | Deep learning; ‘facilitating rapid decisions … in-hospital isolation, treatment facilities, or self-quarantine orders in ED or screening clinics, particularly during pandemics ’ (pp. 8-9); CXRs | C-19; Physicians |
| Jiao, Article – AI development and evaluation | 2020, China (75 patients) | 2, 3 | Deep convolutional neural networks; Automation in breast segmentation and breast mass detection; ‘… potential value for early diagnosis and treatment of breast cancer’ (p. 10)’; MR images | Breast cancer; Radiologists |
| Jing, Article – AI development and evaluation | 2022, The Netherlands (1447 breast MRI examinations of 809 patients) | 2, 2 | Deep learning (breast segmentation, maximum intensive projection generation and abnormality prediction); ‘… workload reduction of 30.6% at the breast level or 15.7% at the examination level’ (p. n.p.)’, ‘… 16.6% to 30.2% of scanner time could be saved over 178 examinations’; ‘excluding normal scans and improving throughput by reducing scanning time … support radiologists’ image interpretation’; MR images | Breast lesions; Radiologists |
| Johansson, Article – AI evaluation; Comparison of AI in radiology against detection by histopathology | 2021, Sweden (120 patient examinations in evaluation) | 3, 2 | Computer-aided detection (CAD)/AI programme (Transpara), deep learning to detect malignant lesions; ‘Transpara … to exclude low-risk cases … radiologists’ workload (may be decreased) by 47% … (however) rate of missed cancer cases … 7%’ (p. 5), best application of Transpara ‘to preselect and exclude low-risk mammographs’ (p. 5); Mammography | Malignant lesions in mammography screening’ Radiologists |
| Joshi, Article – AI development and evaluation | 2021, India, Czech Republic, Italy, Switzerland, Spain (dataset A (publicly available): 237 CXRs; dataset B (publicly available): 5848 CXRs; dataset C (Indian hospitals): 188 + 68 + 543 CXRs) | 3, 3 | Deep learning for automatic diagnosis; ‘pre-screening’, ‘rapid disease detection in 0.137 s per image … to generate probabilistic report in real-time’; CXRs | C-19 detection; Radiologists |
| Kakeda, Article – AI evaluation | 2008, Japan (50 MR angiograms for observer performance study) | 3, 3 | Computer-aided diagnosis (CAD) to shorten reading time yet maintain MRA accuracy; ‘mean reading time of less experienced radiologists with CAD was significantly shorter than that of neuroradiologists without CAD (39.8 vs 54.5 seconds …)’ (p. 459); ‘CAD did not improve the diagnostic accuracy of neuroradiologists …’ (p. 464); MR angiography | Intracranial aneurysms; Radiologists/Neuroradiologists |
| Kasinathan, Article – AI development and evaluation | 2022, India (94 individuals with PET/CT scans; training set: LIDC/IDRI 1024 cases) | 3, 3 | Cloud-based detector and classifier; deep learning; ‘aid better decision-making’; PET/CT images | Classification of lung tumour; Radiologists |
| Keidar, Article – AI development and evaluation | 2021, Switzerland and Israel (1384 CXRs of C-19+, 1024 CXRs of C-19-) | 2, 3 | Deep learning; automated diagnosis and monitoring, ‘aid medical judgment …, improved turnaround times’ (p. 9661); CXRs | C-19 detection; Radiologists |
| Khaled, Article – AI development and evaluation | 2021, Spain (46 cases for evaluation from TCGA-BRCA collection (US)) | 3, 2 | Deep learning, U-Net model, Automated segmentation – otherwise time-consuming manual activity; Dynamic Contrast Enhanced Magnetic Resonance Imaging (DCE-MRI) | Breast lesion segmentation; Radiologists |
| Kim, Article – AI development and evaluation/methodology development (transfer learning) | 2018, UK (AI trained on 11112 images; tested on 100 wrist radiographs) | 3, 3 | Transfer learning from deep convolutional neural networks (CNNs); Automated fracture detection, ‘workflow prioritisation and minimisation of error’ (p. 442); X-rays | Fracture detection; Radiologists |
| Lan, Full conference proceeding – AI development | 2020, China (DDSM (US) database, publicly available, mammography samples: 2620) | 3, 3 | Deep convolutional neural network for computer aided diagnosis (CAD); Mammography | Breast cancer detection and classification; Radiologists |
| Lancaster, Article – AI evaluation (versus radiologist) | 2022, The Netherlands and Russia (283 participants’ ultra-LDCT scans used in evaluation) | 2, 2 | ‘… AI lung cancer screening prototype …’ (p. 134); CAD; Workload reduction by up to 86.7%; ‘AI as an impartial reader in ultra-low-dose CT lung cancer baseline screening ... outperforms all except one experienced radiologist … when looking specifically at negative misclassifications …’ (p. 136), ‘AI … as a first read filter …’ (p. 138); CT scans | Lung nodule detection; Radiologists |
| Lauritzen, Article – AI evaluation | 2022, Denmark (114 421 screenings for breast cancer in 114 421 women) | 2, 2 | AI = Transpara (p. 43); CAD; Radiologist workload reduction by 62.6%, and 25.1% of false-positive screenings; Mammography | National (Denmark) biennial breast cancer screening; Radiologists |
| Li, Full conference proceeding – AI development and evaluation | 2020, China (68 low-dose 3D CT images from pre-op. patient with TAPVC) | 2, 2 | Deep learning for automated segmentation; ‘… improve the efficiency and reduce the workloads of radiologists (400 milliseconds vs. 2-3 hours per-case)’; CT images | Segmentation for total anomalous pulmonary venous connection (TAPVC); Radiologists |
| Li, Article – AI evaluation | 2021, China and US (498 3D images across 2 body regions (head and neck and thorax) IRB from US)) | 2, 2 | SparseGT (AI); automated segmentation; ‘over 90% workload reduction is workload feasible’; CT scans of head and neck and thorax | Automate segmentation with AI; Radiologists |
| Lin, Article – AI development and evaluation | 2022, China (VinDR-CXR – large publicly available dataset) | 3, 3 | Computer-aided diagnosis; Automated detection; CXRs | Multiple lung lesion detection and classification |
| Liu, Article – AI evaluation | 2020, China (590 patients underwent CTPA – training set; 288 patients in validation set) | 2, 3 | Fully deep learning convolutional neural network – U-Net; CTPA (CT pulmonary angiography) | Acute pulmonary embolism detection and quantification of clot burden; Radiologists |
| Liu, Article – AI evaluation | 2021, China (800 ALN lesion samples) | 3, 3 | Traditional machine learning and deep learning to predict ALN metastasis … CECT images … with segmentation; ‘to assist … in determining ALN metastasis …’; CT images | Axiliary lymph node metastasis prediction; Doctors |
| Loizidou, Full conference proceeding – AI development and evaluation | 2021, Cyprus (160 images) | 3, 3 | Temporal subtraction of sequential mammograms, image registration and machine learning; CAD system to ‘assist’ radiologists; Mammography | Automated detection and classification of breast masses; Radiologists |
| Luo, Article – AI development and evaluation | 2022, China (71 patients) | 3, 3 | Convolutional neural network; Automatic detection and segmentation of the thyroid gland; Ultrasonography | Automated detection and segmentation of thyroid gland |
| Ma, Article – AI development and evaluation | 2022, China (‘… millions of natural images’ (p. 366) | 3, 3 | Densely connected convolutional network (SDenseNet and Mul-DenseNet); ‘… reduce heavy workload … avoid misdiagnosis’; Ultrasound | Automatic joint segmentation of thyroid and breast lesions; Physicians |
| Ma, Article – AI development and evaluation | 2017, China (1004 CT cases) | 3, 3 | Radiomics; automatic detection, reduction of error; CT scans | Lung nodules; Radiologists |
| Mahmood, Full conference proceeding – AI development an evaluation | 2021, Australia (60 CT scans for training, testing and validation) | 3, 3 | Deep learning – U-net; Automated segmentation; CT scans | Automatic segmentation of thoracic organs; Radiologists |
| McKinney, Article – AI evaluation | 2020, UK and US (25 856 women) | 3, 3 | Deep learning; AI provided non-inferior performance, Workload reduction of second reader by 88%; Mammography | Automation of stages of national breast screening in UK and application in US; Radiologists |
| Meng, Article – AI development and evaluation | 2019, US (480 radiology reports) | 3, 3 | Machine learning; AI – online decision support system …. identify radiological cases for prompt communication to referring physician | Reporting efficiency; Radiologists |
| Monkam, Article – AI evaluation | 2018, China (publicly accessible database: LIDC/IDRI – 1018 CT scans from 1010 patients) | 3, 3 | CNN models; Reduction false positives; CT images | Lung nodules; Radiologists |

Reviewer 2:

| **Lead author (Surname) and Type of item** | **Date of publication and Country of Research** | **Full text screening code** | **How AI impact radiology services’ workload** | **Notes (Disease and radiology professionals)** |
| --- | --- | --- | --- | --- |
| Noshad, Article – AI development and evaluation | 2021, Iran (219 C-19, 1341 normal images, 1345 viral pneumonia from Qatar University, University of Dhaka, collaborators from Pakistan and Malaysia) | 3, 3 | Deep residual neural network; Assist in initial screening, ‘second opinion … decreased workload … more accurate diagnoses’; CXRs | C-19 detection; Radiologists |
| Pan, Article – AI development and evaluation | 2021, China (3555 MR images) | 3, 2 | Deep convolutional neural networks; Automated diagnosis disc bulge and herniation; ‘improve diagnostic efficiency’; MR images | Lumbar disc bulge, disc herniation; Radiologists |
| Parascandolo, Article - AI development and evaluation | 2015, Italy (57 patients) | 3, 3 | RheumaSCORE: A CAD (via semi-automated segmentation) for rheumatoid arthritis diagnosis; MRI | Rheumatoid arthritis; Radiologists |
| Pedrosa, Article – AI development and evaluation | 2022, Portugal (3 public datasets, 1 private dataset – international) | 3, 3 | Deep learning; Second opinion to support triage of C-19; Assess clinical applicability of deep learning for C-19 screening; CXRs | C-19; Technicians and radiologists |
| Polat, Article – AI development and evaluation | 2021, Turkey (102 CT images for evaluation from 2 publicly available datasets) | 3, 3 | Deep convolutional neural networks; ‘COVID-19 pneumonia … localized automatically … lesion densities can be … evaluated quantitatively’; CT images | C-19 pneumonia; Radiologists |
| Pongsakonpruttikul, Article – AI development and evaluation | 2022, Thailand (1650 knee radiographs – publicly available) | 3, 3 | Deep learning; ‘diagnostic aids’, ‘interpretation and classification of knee OA …’; X-rays | Knee osteoarthritis; Physicians (including radiologists) |
| Postalcioglu, Article – AI development and evaluation | 2022, Turkey (dataset from Kaggle: 2400 CXRs? for training and 600 for testing) | 2, 2 | Boosting techniques/gradient boosting machine learning; Automated diagnosis, ‘pneumonia diagnosis … quickly and accurately by anyone using X-ray’ (p. 14); CXRs | Pneumonia (C-19); Doctors/radiologists |
| Prasad, Full conference proceeding – AI development and evaluation | 2022, India (C-19 dataset collection from Kaggle, CXRs | 3, 3 | CNN, deep learning; ‘diagnostic decision support device’; CXRs | C-19 detection; Radiologist |
| Purkayastha, Article – AI development and evaluation | 2020, US (>100 000 CXRs from publicly available dataset - CheXNet) | 3, 3 | Neural network algorithm; ‘turnaround time of under 30 s’; ‘feasibility of a web service for machine learning based diagnosis of 14 lung pathologies’ (p. 1), Aim for implementation in areas of few radiologists; CXRs | Thoracic diseases; Radiologists |
| Qi, Article – AI development and evaluation | 2021, China (130 NPC patients – CT; 149 NPC patients – MR images) | 3, 3 | Computer-aided diagnosis and regional segmentation for treatment; Automation of diagnosis and treatment planning; CT and MR images | Nasopharyngeal carcinoma; Radiologists |
| Qiu, Full conference proceeding – AI development and evaluation | 2018, China (312 patients) | 3, 3 | Texture analysis method/radiomics; ‘… reference for diagnosis … reference value for the quantitative data research … pancreatic lesions …’ (p. 12); CT images | Pancreatic cancer; Radiologists |
| Ragab, Article – AI development and evaluation | 2020, Egypt (SARS-CoV-2 CT-scan dataset from Brazil: 1252 CT images/60 patients and 1230 CT images/60 patients) | 3, 2 | FUSI-CAD, convolutional neural networks; ‘proposed system …. simple to set up, low cost, and automated CAD … accurate, effective, and fast diagnostic tool … early diagnosis’ (p. 20); CT images | C-19 diagnosis; Radiologists |
| Rajaraman, Article – AI development and evaluation | 2021, US (Multiple international datasets see pp. 5-7) | 3, 3 | U-Net models for semantic segmentation; ‘TB classification and segmentation’ (p. 27), ‘advanced assistance in radiologist interpretive workflows: … triage … classification … improving productivity ...’ (p. 28); CXRs | Tuberculosis; Radiologists |
| Rao, Article – AI application and evaluation | 2021, US (6565 non-contrast brain CT scans) | 2, 3 | AI ‘adjunct to current peer review tools as a second reader …’ (p. 92); CT images | Intracranial haemorrhage; Radiologists |
| Raya-Povedano, Article – AI development and evaluation | 2021, Spain (15 987 digital mammography and digital breast tomosynthesis examinations) | 2, 2 | AI/Transpara with DBT would result in 72.5% less workload and AI with DM would result in 29.7%; ‘… screening workload could be safely reduced up to 70% for both digital mammography (DM) – and digital breast tomosynthesis (DBT) …’ (p. 64); DM and DBT images | National breast screening programme; Radiologists |
| Rodriguez-Ruiz, Article – AI application and evaluation | 2019, The Netherlands, Switzerland, Italy, Austria, Spain, Germany and Sweden (2652/2654? DM examinations) | 2, 2 | AI/CAD for triage; AI mid-point score … halving workload …; Mammography | Breast screening programme |
| Saba, Article – AI development and evaluation | 2022, Saudi Arabia (780 images) | 2, 3 | CAD, convolutional neural networks; ‘CAD … help … identify malignant cases with minimal time and effort …’ (p. 7); Breast ultrasound images | Breast ultrasound; Radiologists |
| Salem Salamh, Article – AI development and evaluation | 2021, Turkey (800 images in development) | 2, 3 | Convolutional neural network, VGG/GK-Tool; ‘good recognition rates in the diagnosis … could reduce the radiologist’s workload’ (p. 1); CT images | C-19 detection and classification; Radiologists |
| Seidel, Full conference proceeding – AI development and evaluation | 2014, US (NCI Lung Image Database Consortium (LIDC) dataset) | 2, 2 | CAD; ‘… metric-based case partitioning can be used to better select … radiologists assigned to each case … further assist with diagnosis … to shed 25% of radiologist annotations without any loss of predictive accuracy’ (p. 771); CT | Diagnostic consensus in image interpretation; Radiologists |
| Sheela, Article – AI development and evaluation | 2022, India (200 training cases/MR images) | 2, 2 | AI algorithm; ‘…classify the presence of pneumonia which will in turn save around 50% of the time frame for physicians …’ (p. 2049); MR images | C-19 detection, early stage |
| Shibata, Article – AI development and evaluation | 2021, Japan (3845 subjects non-contrast head CTs) | 2, 2 | Semi-supervised flow-based generative models for ‘versatile anomaly detection method’ (p. 1), ‘reduce the workload for labeling’ (p. 5); CXRs and brain CTs (BCTs) | Lesion detection and qualification; Radiologists |
| Shimada, Article – AI development and evaluation | 2020, Japan (1623 subjects, 5 subjects with missed aneurysms initially) | 3, 3 | CAD using convolutional neural network; ‘CAD … might pave the way to substitute the workload of diagnostic radiologists and reduce the cost of human labor’ (p. 4); MR images | Cerebral aneurysms; Radiologists |
| Shoshan, Article – AI development and evaluation | 2022, US (13 306 DBT examinations/9919 women; model tested on 4310 screened women) | 2, 2 | ‘… use of AI to automatically filter out cases would result in 39.6% less workload … 25% lower recall rate’; digital breast tomosynthesis (DBT) | Breast screening; Radiologists |
| Sichtermann, Article – AI development and evaluation | 2019, Germany (85 examinations) | 3, 2 | Deep learning convolutional neural network; Automated detection (CAD) of intracranial aneurysm (IA), ‘potential for reliable detection of (IA) from 3D TOF-MRA’ (p. 30), ‘(CAD) … preventing diagnostics errors … physician’s fatigue’ (p. 30) | Intracranial aneurysms; Radiologists |
| Siddiqui, Article – AI development and evaluation | 2020, Pakistan (527 images) | 3, 3 | Deep learning, convolutional neural network; CT scans and X-rays | C-19 detection; Radiologists and medical practitioners |
| Su, Article – AI development and evaluation | 2021, China (1018 patients) | 2, 2 | Convolutional neural network; Automated lung nodule detection; ‘reduces … rate of misdiagnosis and missed diagnosis’; CT images | Lung nodule detection; Radiologists |
| Tandungan, Full conference proceeding - AI development and evaluation | 2019, Indonesia (use of LIDC-IDRI dataset) | 3, 3 | CAD/Extreme Learning Machine for automated diagnosis, ‘help radiologists in analyzing lung cancer nodules …’ (p. 1); CT scans | Lung cancer classification |
| Tsai, Full conference proceeding – AI development and evaluation | 2012, Taiwan (4 datasets – no additional information) | 2, 3 | CAD/Multiple Active Contour Models and Gabor Neural Network; Diagnostic accuracy; CT | Pulmonary embolism; No clinician group identified |
| Tsai, Article – AI development and evaluation | 2022, Taiwan (5733 mammograms/1490 patients) | 2, 3 | Deep neural network; Mammographic interpretation; Mammograms | Breast screening; Radiologists |
| Van den Oever, Article – AI development and evaluation | 2020, The Netherlands (60 scans) | 3, 2 | Deep learning; Automated detection/exclusion or segmentation on cardiac CT of coronary artery calcium; cardiac CT images | Coronary artery calcium (CAC) detection; Radiologist |
| Verburg, Article – AI development and evaluation | 2022, The Netherlands (4581 breast examinations) | 3, 2 | Deep learning; Automated triaging to differentiate breasts with lesions and those without … to dismiss from radiologic review; MRI examinations | Breast screening; Radiologists |
| Vilares, Full conference proceeding – AI development and evaluation | 2021, Portugal (259 mammograms with lesions, 255 normal) | 3, 2 | Deep learning; Triage complex from normal to enable radiologists to focus on complex; Mammograms | Breast screening; Radiologists |
| Wang, Article – AI development and evaluation | 2018, China (2480 images from public datasets) | 3, 3 | Deep learning; ‘… provide physicians and radiologists with valuable information to significantly decrease time-to-diagnosis’ (p. 16); CXRs | Lung lesions; Radiologists |
| Wang, Article – AI development and evaluation | 2021, China (25000 scans to develop model; validation 75 and 491 scans) | 3, 3 | Deep learning, 2D CNN model; Automated detection and classification of AIH; ‘prompt and better decision-making’ (p. 1), ‘second-read or triage tool’ (p. 1); CT scans | Acute intracranial haemorrhage; Radiologists |
| Wong, Full conference presentation – AI development and evaluation | 2019, US (>100 000 images from 30 000 patients from NIH Clinical Center) | 2, 2 | ‘Deep learning framework (CNN model) for normal/abnormal classification (of chest X-rays)’ (p. 6); ‘average recall of 50% … cut in half the number of disease-free CXRs examined by radiologists’ (p. 1); CXRs | Disease-free CXRs; Radiologists |
| Wu, Article – AI development and evaluation | 2020, China (495 patients from 3 hospitals) | 3, 3 | Deep learning; ‘improve the efficacy of diagnosis’ (p. 1); CT images | C-19 screening; Radiologists |
| Yala, Article – AI development and evaluation | 2019, US (223 109 screening mammograms of 66 661 women 2009 – 2016) | 2, 2 | Deep learning to triage/pre-selecting mammograms; ‘cancer free’ not reviewed by radiologist; ‘workload reduction of 19.3%’ (p. 43) | Screening mammograms; Radiologist |
| Yan, Article – AI development and evaluation | 2020, China (206 C-19 patients with 416 CT scans and 412 CP patients with 412 CT scans) | 2, 2 | Deep learning; multi-scale convolutional neural network (MSCNN); ‘rapid diagnosis’; CT scans | C-19 distinction from common pneumonia; Radiologists and physicians  EXCLUDE? – minimal detail re: impact on radiologist/physician workload beyond ‘quick diagnosis’ |
| Yang, Article – AI development and evaluation | 2020, China (295 patients) | 2, 3 | Deep learning; DenseNet; Improved diagnostic efficiency; CT scan | C-19 detection; Radiologists |
| Yang, Article – AI development and evaluation | 2021, China (2749 cases) | 3, 3 | AI-powered mammographic breast lesion diagnostic system; Accelerate diagnostic process; Mammography | Mammography screening; Radiologists |
| Yao, Article – AI development and evaluation | 2021, China (1707 patients) | 3, 3 | Deep learning; ‘superior recall and similar diagnostic precision to (experienced) radiologists’ (p. 8); CT scans | Rib fracture detection; Radiologists |
| Yates, Article – AI development and evaluation | 2018, UK (ChestX-ray14 from NIH: 112 120 images from 30 000 patients and 7470 from Indiana University hospital network chest radiograph database) | 3, 3 | Open-source, cloud, deep convolutional neural networks; ‘classify chest radiographs as normal or abnormal’; CXRs | Chest radiograph classification; Radiologists |
| Zamacona, Article – AI development and evaluation | 2015, US (LIDC dataset) | 2, 3 | CAD; Categorising diagnostic complexity, ‘determine the easy from hard …’; ‘… best allocate additional radiologists to interpret a case based on its diagnostic category’; CT scans | Lung nodule image dataset used for AI development; Radiologists |
| Zapaishchykova, Article – AI development and evaluation | 2021, US (373 admission CT scans from 2 level 1 trauma centres) | 2, 3 | Faster-RCNN; automation; ‘prioritize the reading queue of the attending trauma radiologist’; ‘extract pelvic-fracture-related risk scores’ (p. 425); CT scans | Pelvic trauma severity scoring; Radiologists |
| Zhang, Article – AI development and evaluation | 2019, China (18 cases, 8 with lung ca. and 10 without) | 2, 3 | Computer-aided diagnosis, Multiscale Mask R-CNN; ‘effectiveness of method in detecting lung tumors along with the capability of identifying a healthy chest pattern and reducing incorrect identification of tumors …’(p. 1); PET imaging | Lung tumour detection |
| Zhang, Article - AI development and evaluation | 2021, China (416 C-19 CT scans from 216 patients, 412 CP CT scans from 412 patients) | 2, 2 | U-Net and ResNet-18; Segmentation and classification; ‘assisting physicians and radiologists in rapid COVID-19 detection’; CT images | C-19 detection |
| Zhao, Full conference proceeding – AI development and evaluation | 2022, China (400 patient with confirmed glioma) | 3, 3 | CAD, Radiomics; ‘reduce time of manual segmentation’; MR images | Glioma grading; Radiologists |
| Zhou, Article – AI development and evaluation | 2020, China (1079 patients) | 2, 3 | Faster R-CNN; Automatic detection and classification; CT scans | Rib fractures on thoracic CT scans; Radiologists |

Notes

Full text screening codes:

- ‘2’s about volume of images
- ‘3’s about efficiency

Exclusions:

- NA – full paper/article not available: 23
- NE – not in English: 2
- Review - 3: Ahmad (2021); Sechopoulos and Mann (2020); Shi et al. (2020)
- Conference abstract only – 6: Hernandez et al. (2021); Raghav et al. (2020); Tan and Parizel (2021); Tomori et al. (2018); Vonder et al. (2021); Yuan et al. (2018)
- AI for other staff groups beyond clinical radiology services with diagnostic remit – 5: for example, radiologists and dentists (Chen et al. 2020); physician workload (Clymer et al. 2020); orthopaedic surgeons (Li et al. 2019); obstetricians (Liu et al. 20220); radiologists and neurosurgeons (Shi et al. 2020)
- Radiotherapy - 3: Choi et al. (2020); Huang et al. (2021); Olsen et al. (2012)
- AI for treatment follow-up – 4: El Adoui et al. (2019); Sullivan et al. (2020); Zhao et al. (2020); Zhao et al. (2022)
- AI during surgery – 2: surgery navigation (Hu et al. 2022); surgical planning (Li et al. 2021)
- AI development – 10: Park et al. (2021); Peng et al. (2021); Petrov et al. (2017); Shu et al. (2020); Sotoudeh-Paima et al. (2022); Sun et al. (20221)/Peng et al. (2021); Tamer et al. (2022); Yeh et al. (2018); Yu et al. (2020); Yu et al. (2021)
- AI for communication/reporting – 3: Spandorfer et al. (2019); Yuan et al. (2019); Zhang et al. (2020)
- Evaluation of AI with limited explanation of impact on radiology workload – 5: Lin et al. (2021); Taylor et al. (2008); Vorontsov et al. (2019); Yang et al. (2021) – ‘Integrate domain knowledge …’; Zeng et al. (2021)
- Other technology – 7: Miles et al. (2018); Mizan et al. (2021); Mohsen et al. (2014); Patil et al. (2016); Ritchie et al. (2016); Taylor (2015); Vasilakakis et al. (2019)
- Other – 2: Van den Biggelaar et al. (2009) – use of breast technologists in pre-reading mammograms; Yamaguchi et al. (2020) – study protocol

**Appendix C Quality appraisal of included studies using the Mixed Methods Appraisal Tool^[1]^ (2018)**

| **Included studies** | **Quantitative non-randomized** | | | | | **Quantitative descriptive** | | | | | **Mixed methods** | | | | |
| --- | --- | --- | --- | --- | --- | --- | --- | --- | --- | --- | --- | --- | --- | --- | --- |
|  | **3.1** | **3.2** | **3.3** | **3.4** | **3.5** | **4.1** | **4.2** | **4.3** | **4.4** | **4.5** | **5.1** | **5.2** | **5.3** | **5.4** | **5.5** |
| Abbas et al. (2020) | - | - | - | - | - | Y | Y | Y | NA | ? | - | - | - | - | - |
| Agrawal and Choudhary (2022) | - | - | - | - | - | Y | Y | Y | NA | Y | - | - | - | - | - |
| Aiello et al. (2022) | Y | Y | Y | ? | Y | - | - | - | - | - | - | - | - | - | - |
| Alajmi et al. (2022) | - | - | - | - | - | Y | Y | Y | NA | Y | - | - | - | - | - |
| Alduraibi (2022) | - | - | - | - | - | Y | Y | Y | NA | Y | - | - | - | - | - |
| Ali et al. (2020) | - | - | - | - | - | Y | Y | Y | NA | ? | - | - | - | - | - |
| Allaouzi et al. (2021) | - | - | - | - | - | Y | Y | Y | NA | Y | - | - | - | - | - |
| Amara et al. (2022) | Y | Y | Y | ? | Y | Y | Y | Y | NA | y | - | - | - | - | - |
| Antonios et al. (2013) | - | - | - | - | - | Y | Y | Y | NA | y | - | - | - | - | - |
| Ardakani et al. (2020) | - | - | - | - | - | Y | Y | Y | NA | y | - | - | - | - | - |
| Benedikt et al. (2018) | Y | Y | Y | ? | Y | - | - | - | - | - | - | - | - | - | - |
| Calisto et al. (2021) | - | - | - | - | - | - | - | - | - | - | Y | Y | Y | Y | Y |
| Cao et al. (2022) | Y | Y | Y | ? | Y | - | - | - | - | - | - | - | - | - | - |
| Chang et al. (2022) | - | - | - | - | - | Y | Y | Y | NA | Y | - | - | - | - | - |
| Chen et al. (2021a) | - | - | - | - | - | Y | Y | Y | NA | Y | - | - | - | - | - |
| Chen et al. (2021b) | - | - | - | - | - | Y | Y | Y | NA | ? | - | - | - | - | - |
| Clymer et al. (2020) | - | - | - | - | - | Y | Y | Y | NA | ? | - | - | - | - | - |
| Cunha et al. (2020) | Y | Y | Y | ? | Y | - | - | - | - | - | - | - | - | - | - |
| Dai et al. (2021) | Y | Y | Y | ? | Y | - | - | - | - | - | - | - | - | - | - |
| Dembrower et al. (2020) | Y | Y | Y | ? | Y | - | - | - | - | - | - | - | - | - | - |
| Duron et al. (2022) | - | - | - | - | - | Y | Y | Y | NA | Y | - | - | - | - | - |
| Dyer et al. (2022) | - | - | - | - | - | Y | Y | Y | NA | Y | - | - | - | - | - |
| Dyer et al. (2021) | - | - | - | - | - | Y | Y | Y | NA | Y | - | - | - | - | - |
| Feng et al. (2021) | - | - | - | - | - | Y | Y | Y | NA | ? | - | - | - | - | - |
| Gaal et al. (2020) | - | - | - | - | - | Y | Y | Y | NA | ? | - | - | - | - | - |
| Galvan-Tejada et al. (2017) | Y | Y | Y | ? | Y | - | - | - | - | - | - | - | - | - | - |
| Ghosal et al. (2021) | - | - | - | - | - | Y | Y | Y | NA | ? | - | - | - | - | - |
| Grauhan et al. (2022) | Y | Y | Y | ? | Y | - | - | - | - | - | - | - | - | - | - |
| Hallinan et al. (2021) | Y | Y | Y | ? | Y | - | - | - | - | - | - | - | - | - | - |
| Han et al. (2022) | - | - | - | - | - | Y | Y | Y | NA | ? | - | - | - | - | - |
| Hidayah et al. (2015) | - | - | - | - | - | Y | Y | Y | NA | ? | - | - | - | - | - |
| Hsieh et al. (2021) | - | - | - | - | - | Y | Y | Y | NA | ? | - | - | - | - | - |
| Huang et al. (2019) | - | - | - | - | - | Y | Y | Y | NA | ? | - | - | - | - | - |
| Hussain and Ruza (2022) | - | - | - | - | - | Y | Y | Y | NA | ? | - | - | - | - | - |
| Ismaeil and Salem (2020) | - | - | - | - | - | Y | Y | Y | NA | ? | - | - | - | - | - |
| Jacob et al. (2021) | - | - | - | - | - | Y | Y | Y | NA | ? | - | - | - | - | - |
| Jang et al. (2020) | Y | Y | Y | ? | Y | - | - | - | - | - | - | - | - | - | - |
| Jiao et al. (2020) | Y | Y | Y | ? | Y | - | - | - | - | - | - | - | - | - | - |
| Jing et al. (2022) | Y | Y | Y | ? | Y | - | - | - | - | - | - | - | - | - | - |
| Johansson et al. (2021) | Y | Y | Y | ? | Y | - | - | - | - | - | - | - | - | - | - |
| Joshi et al. (2021) | - | - | - | - | - | Y | Y | Y | NA | ? | - | - | - | - | - |
| Kakeda et al. (2008) | Y | Y | Y | ? | Y | - | - | - | - | - | - | - | - | - | - |
| Kasinathan and Jayakumar (2022) | Y | Y | Y | ? | Y | - | - | - | - | - | - | - | - | - | - |
| Keidar et al. (2021) | - | - | - | - | - | Y | Y | Y | NA | Y | - | - | - | - | - |
| Khaled et al. (2022) | - | - | - | - | - | Y | Y | Y | NA | ? | - | - | - | - | - |
| Kim and MacKinnon (2018) | - | - | - | - | - | Y | Y | Y | NA | Y | - | - | - | - | - |
| Lan and Zhong (2020) | - | - | - | - | - | Y | Y | Y | NA | ? | - | - | - | - | - |
| Lancaster et al. (2022) | Y | Y | Y | ? | Y | - | - | - | - | - | - | - | - | - | - |
| Lauritzen et al. (2022) | Y | Y | Y | ? | Y | - | - | - | - | - | - | - | - | - | - |
| Li et al. (2020) | - | - | - | - | - | Y | Y | Y | NA | ? | - | - | - | - | - |
| Li et al. (2021) | - | - | - | - | - | Y | Y | Y | NA | ? | - | - | - | - | - |
| Lin et al. (2022) | - | - | - | - | - | Y | Y | Y | NA | ? | - | - | - | - | - |
| Liu et al. (2020) | Y | Y | Y | ? | Y | - | - | - | - | - | - | - | - | - | - |
| Liu et al. (2021) | - | - | - | - | - | Y | Y | Y | NA | ? | - | - | - | - | - |
| Loizidou et al. (2021) | - | - | - | - | - | Y | Y | Y | NA | ? | - | - | - | - | - |
| Luo et al. (2022) | Y | Y | Y | ? | Y | - | - | - | - | - | - | - | - | - | - |
| Ma et al. (2022) | - | - | - | - | - | Y | Y | Y | NA | ? | - | - | - | - | - |
| Ma et al. (2017) | - | - | - | - | - | Y | Y | Y | NA | ? | - | - | - | - | - |
| Mahmood et al. (2021) | - | - | - | - | - | Y | Y | Y | NA | ? | - | - | - | - | - |
| McKinney et al. (2020) | - | - | - | - | - | Y | Y | Y | NA | ? | - | - | - | - | - |
| Meng et al. (2019) | - | - | - | - | - | Y | Y | Y | NA | ? | - | - | - | - | - |
| Monkam et al. (2018) | - | - | - | - | - | Y | Y | Y | NA | ? | - | - | - | - | - |
| Noshad et al. (2021) | - | - | - | - | - | Y | Y | Y | NA | ? | - | - | - | - | - |
| Pan et al. (2021) | - | - | - | - | - | Y | Y | Y | NA | ? | - | - | - | - | - |
| Parascandolo et al. (2015) | Y | Y | ? | ? | ? | - | - | - | - | - | - | - | - | - | - |
| Pedrosa et al. (2022) | - | - | - | - | - | Y | Y | Y | NA | ? | - | - | - | - | - |
| Polat et al. (2021) | - | - | - | - | - | Y | Y | Y | NA | ? | - | - | - | - | - |
| Pongsakonpruttikul et al. (2022) | - | - | - | - | - | Y | Y | Y | NA | ? | - | - | - | - | - |
| Postalcioglu (2022) | - | - | - | - | - | Y | Y | Y | NA | ? | - | - | - | - | - |
| Prasad et al. (2022) | - | - | - | - | - | Y | Y | Y | NA | ? | - | - | - | - | - |
| Purkayastha et al. (2020) | - | - | - | - | - | Y | Y | Y | NA | ? | - | - | - | - | - |
| Qi et al. (2021) | Y | Y | Y | ? | Y | - | - | - | - | - | - | - | - | - | - |
| Qiu et al. (2018) | - | - | - | - | - | Y | Y | Y | NA | ? | - | - | - | - | - |
| Ragab et al. (2020) | - | - | - | - | - | Y | Y | Y | NA | ? | - | - | - | - | - |
| Rajaraman et al. (2021) | - | - | - | - | - | Y | Y | Y | NA | Y | - | - | - | - | - |
| Rao et al. (2021) | - | - | - | - | - | Y | Y | Y | NA | ? | - | - | - | - | - |
| Raya-Povedano et al. (2021) | - | - | - | - | - | Y | Y | Y | NA | Y | - | - | - | - | - |
| Rodriguez-Ruiz et al. (2019) | - | - | - | - | - | Y | Y | Y | NA | Y | - | - | - | - | - |
| Saba et al. (2022) | - | - | - | - | - | Y | Y | Y | NA | ? | - | - | - | - | - |
| Salem Salamh et al. (2021) | - | - | - | - | - | Y | Y | Y | NA | ? | - | - | - | - | - |
| Seidel et al. (2014) | - | - | - | - | - | Y | Y | Y | NA | ? | - | - | - | - | - |
| Sheela and Arun (2022) | - | - | - | - | - | Y | Y | Y | NA | ? | - | - | - | - | - |
| Shibata et al. (2021) | Y | Y | Y | ? | Y | - | - | - | - | - | - | - | - | - | - |
| Shimada et al. (2020) | Y | Y | Y | ? | Y | - | - | - | - | - | - | - | - | - | - |
| Shoshan et al. (2022) | - | - | - | - | - | Y | Y | Y | NA | Y | - | - | - | - | - |
| Sichtermann et al. (2019) | Y | Y | Y | ? | Y | - | - | - | - | - | - | - | - | - | - |
| Siddiqui et al. (2020) | - | - | - | - | - | Y | Y | Y | NA | ? | - | - | - | - | - |
| Su et al. (2021) | - | - | - | - | - | Y | Y | Y | NA | ? | - | - | - | - | - |
| Tandungan et al. (2019) | - | - | - | - | - | Y | Y | Y | NA | ? | - | - | - | - | - |
| Tsai et al. (2012) | - | - | - | - | - | Y | Y | Y | NA | ? | - | - | - | - | - |
| Tsai et al. (2022) | - | - | - | - | - | Y | Y | Y | NA | ? | - | - | - | - | - |
| Van den Oever et al. (2020) | Y | Y | Y | ? | Y | - | - | - | - | - | - | - | - | - | - |
| Verburg et al. (2022) | Y | Y | Y | ? | Y | - | - | - | - | - | - | - | - | - | - |
| Vilares et al. (2021) | - | - | - | - | - | Y | Y | Y | NA | ? | - | - | - | - | - |
| Wang et al. (2018) | - | - | - | - | - | Y | Y | Y | NA | ? | - | - | - | - | - |
| Wang et al. (2021) | - | - | - | - | - | Y | Y | Y | NA | Y | - | - | - | - | - |
| Wong et al. (2019) | - | - | - | - | - | Y | Y | Y | NA | ? | - | - | - | - | - |
| Wu et al. (2020) | Y | Y | Y | ? | Y | - | - | - | - | - | - | - | - | - | - |
| Yala et al. (2019) | Y | Y | Y | ? | Y | - | - | - | - | - | - | - | - | - | - |
| Yan et al. (2020) | Y | Y | Y | ? | Y | - | - | - | - | - | - | - | - | - | - |
| Yang et al. (2020) | Y | Y | Y | ? | Y | - | - | - | - | - | - | - | - | - | - |
| Yang et al. (2021) | - | - | - | - | - | Y | Y | Y | NA | Y | - | - | - | - | - |
| Yao et al. (2021) | Y | Y | Y | ? | Y | - | - | - | - | - | - | - | - | - | - |
| Yates et al. (2018) | - | - | - | - | - | Y | Y | Y | NA | Y | - | - | - | - | - |
| Zamacona et al. (2015) | - | - | - | - | - | Y | Y | Y | NA | Y | - | - | - | - | - |
| Zapaishchykova et al. (2021) | - | - | - | - | - | Y | Y | Y | NA | ? | - | - | - | - | - |
| Zhang et al. (2019) | - | - | - | - | - | Y | Y | Y | NA | Y | - | - | - | - | - |
| Zhang and Zhang (2021) | - | - | - | - | - | Y | Y | Y | NA | ? | - | - | - | - | - |
| Zhao et al. (2022) | - | - | - | - | - | Y | Y | Y | NA | Y | - | - | - | - | - |
| Zhou et al. (2020) | Y | Y | Y | ? | Y | - | - | - | - | - | - | - | - | - | - |

Notes

- No studies were solely qualitative (1. of the MMAT algorithm) or randomised controlled trials (2. of the MMAT algorithm) hence these groups of screening questions have not been incorporated on the above table
- Quantitative non-randomized appraisal criteria: 3.1 Are the participants representative of the target population? 3.2 Are measurements appropriate regarding both the outcome and intervention (or exposure)? 3.3 Are there complete outcome data? 3.4 Are the confounders accounted for in the design and analysis? 3.5 During the study period, is the intervention administered (or exposure) as intended?
- Quantitative descriptive appraisal criteria: 4.1 Is the sampling strategy relevant to address the research question? 4.2 Is the sample representative of the target population? 4.3 Are the measurements appropriate? Is the risk of nonresponse bias low? Is the statistical analysis appropriate to answer the research question?
- Mixed methods appraisal criteria: 5.1 Is there an adequate rationale for using a mixed methods design to address the research question? 5.2 Are the different components of the study effectively integrated to answer the research question? 5.3 Are the outputs of the integration of qualitative and quantitative results adequately addressed? 5.4 Are divergences and inconsistences between quantitative and qualitative results adequately addressed? 5.5 Do the different components of the study adhere to the quality criteria of each tradition of the methods involved?
- Y = yes, the study meets the stated criterion
- N = no, the study does not meet the stated criterion
- NA = not applicable
- ? = can’t tell if the study does or does not meet the stated criterion
- - = questions not used to screen

1. Hong, Q. N., Pluye, P., Fabregues, S., Bartlett, G., Boardman, F., Cargo, M., Dagenais, P., Gagnon, M-P., Griffiths, F., Nicolau, B., O’Cathian, A., Rousseau, M-C., Vedel, I. (2018) *Mixed Methods Appraisal Tool (MMAT) Version 2018, User guide*. http://mixedmethodsappraisaltoolpublic.pbworks.com/w/file/fetch/127916259/MMAT_2018_criteria-manual_2018-08-01_ENG.pdf Accessed 29 July 2022
